# Clinical Validation of a Quantitative HIV-1 DNA Droplet Digital PCR assay: Applications for Detecting Occult HIV-1 Infection and Monitoring Cell-associated HIV-1 Dynamics across Different Subtypes in HIV-1 Prevention and Cure Trials

**DOI:** 10.1101/2020.09.29.20200154

**Authors:** Laura Powell, Adit Dhummakupt, Lilly Siems, Dolly Singh, Yann Le Duff, Priyanka Uprety, Cheryl Jennings, Joseph Szewczyk, Ya Chen, Eleni Nastouli, Deborah Persaud

**Affiliations:** Johns Hopkins University, School of Medicine, Department of Pediatrics, Division of Infectious Diseases, Baltimore, MD; Center for AIDS reagents, National Institute for Biological Standards and Controls, England, UK; Department of Pathology and Laboratory Medicine, Robert Wood Johnson University Hospital, Rutgers University, New Brunswick, NJ; Rush University Medical Center, Department of Molecular Pathogens and Immunity, Chicago, IL; Department of Population, Policy and Practice, UCL Great Ormond Street Institute of Child Health and Francis Crick Institute, London, UK; Departments of Molecular Microbiology and Immunology and International Health, Johns Hopkins Bloomberg School of Public Health

## Abstract

**Background:** In HIV-1-exposed infants, nucleic acid testing (NAT) is required to diagnose infection since passively transferred maternal antibodies preclude antibody testing. The sensitivity of clinical NAT assays is lowered with infant antiretroviral prophylaxis and, with empiric very early antiretroviral treatment of high-risk infants, thereby impacting early infant diagnosis. Similarly, adult HIV-1 infections acquired under pre-exposure prophylaxis may occur at low levels (occult infection), with undetectable plasma viremia and indeterminate antibody tests, for which HIV-1 DNA testing maybe a useful adjunct. Cell-associated HIV-1 DNA concentrations are also used to monitor HIV-1 persistence in viral reservoirs with relevance to HIV-1 cure therapeutics, particularly in perinatal infections.

**Methods:** The analytical sensitivity and specificity of an HIV-1 DNA droplet digital PCR (ddPCR) assay was determined, across different HIV-1 subtypes, using serial dilutions of a plasmid containing a 160-base pair sequence of the HIV-1 LTR-gag spiked into peripheral blood mononuclear cells (PBMCs), with MOLT-4 cells or PBMCs infected with different HIV-1 subtypes (A, B and C), and U1 cells spiked into PBMCs. Inter- and intra-run variability were used to determine assay precision.

**Results:** The HIV-1 LTR-gag ddPCR assay was reliable and reproducible, and exhibited high analytical specificity with sensitivity to near single copy level, across multiple HIV-1 subtypes, and a limit of detection of 4.09 copies/million PBMCs.

**Conclusions:** This assay has applications for detecting occult HIV-1-infection that may occur in the setting of combination and long-acting regimens used for HIV-1 prevention, across different HIV-1 subtypes, in both infants and adults, and in HIV-1 cure interventions, particularly with perinatal infections.

## Introduction

In HIV-1-exposed infants younger than age 18 months, HIV-1 antibody testing cannot distinguish between infant and maternally-derived antibodies to allow use for infant HIV-1 diagnosis (1, 2). Quantitative or qualitative real-time polymerase chain reaction (RT-PCR) assays that detect HIV-1 RNA in plasma or DNA in peripheral blood mononuclear cells (PBMCs) are therefore required (3, 4). Early antiretroviral treatment (ART) of perinatal HIV-1 infection can lead to low concentrations of circulating HIV-1 infected cells in association with negative HIV-1 antibody and DNA tests by clinical diagnostic assays (5-11), which can lead to unnecessary ART interruption in later childhood (12). Furthermore, the use of combination antiretroviral (ARV) prophylaxis in high-risk perinatal exposures, along with very early ART within days of age, or directly transitioning from prophylaxis to ART may lead to low infected cell concentrations (7, 13-16) that require more sensitive HIV-1 DNA assays to confirm infection on ART. HIV-1 DNA concentrations in peripheral blood are also used to determine therapeutic efficacy of cure strategies aimed at eradicating reservoir cells in for ART-free remission and cure (17, 18). Sensitive and clinically validated HIV-1 DNA assays may become useful in these specialized settings, in both perinatal and adult infections. We optimized and clinically validated an HIV-1 DNA quantitative assay using the more sensitive and precise approach of droplet digital PCR (ddPCR), across different HIV-1 subtypes, with potential applications for use clinically and in HIV-1 prevention and cure trials (19).

## Materials and Methods

### A detailed description of the DNA isolation and ddPCR is provided in the supplemental materials

#### Preparation of cell pellets and plasmid standards

PBMCs were isolated from leukopaks (New York City Blood Center, New York NY) using Ficoll-Hypaque centrifugation (GE Healthcare, Chicago IL), treated with Red Blood Cell Lysis Buffer (eBiosciences, San Diego CA) and aliquoted into 5-million cell pellets for storage at −80°C until further use.

#### Plasmid standards

A plasmid comprised of a pMK-RQ-Bs backbone (GeneArt Gene Synthesis Technology; Thermo Scientific, Waltham MA) with a 160-bp region of the HIV-1-5’ LTR and gag region (positions 626 – 786, HXB2) (20), as well as a fragment of the housekeeping gene, Ribonuclease Protein Subunit P30 (RPP30) (21), was constructed (Supplemental Figure 1). Prior to testing, an aliquot of plasmid stock was serially diluted and spiked into 5-million PBMC pellets to reach final analyte concentrations ranging from 1,000 to 1.25 copies per 10^6^ PBMCs. A second high-dilution series (10, 000 to 100 copies per 10^6^ PBMCs) was tested to assess linearity at the higher concentrations.

#### DNA Isolation

Genomic DNA was isolated from the combined plasmid-cell mixtures using QIAamp DNA Blood Midi Kit (Qiagen, Germantown MD) with a modification that included addition of an overnight ethanol precipitation step to obtain genomic DNA of high concentration and purity for downstream ddPCR applications (21).

#### Primers, Probes and PCR conditions

The primers and probes for HIV-1 LTR-gag (20) and RPP30 (21) (Supplemental Table 1) were obtained from Integrated DNA Technologies, Coralville IA), reconstituted in DNase/RNase-free water, and stored at concentrations of 18 µM primers and 5 µM probes in 50 µL aliquots for single use only.

#### Genomic DNA preparation and ddPCR

Genomic DNA (sufficient to perform 8 replicates at 1000ng input per well) was subjected to restriction digest with XbaI (New England BioLabs, Ipswich MA). Negative control wells with genomic DNA without plasmid standards were done in four replicates, along with a single control well without genomic DNA as a no template control (NTC). To directly quantify the number of cells analyzed, based on copies of RPP30, PBMC genomic DNA on each sample was tested in a 1:10 dilution (100 ng DNA per well) since RPP30 is present in high copy number at two copies per cell. This allowed the results to be expressed as HIV-1 DNA copies per cells analyzed with subsequent conversion to copies per 10^6^ cells.

The reaction mixtures of primers, probes, Supermix, sample, and Droplet Generation Oil were placed in the QX-200 Droplet Generator according to manufacturer instructions for droplet generation. PCR reactions were carried out at the conditions summarized in the Supplemental Methods and analyzed on the QX200 Droplet Digital PCR Reader (Bio-Rad, Hercules CA) that assesses individual droplets for their level of fluorescence.

#### Analysis of ddPCR data

Analysis of HIV-1 DNA copies per sample was completed in QuantaSoft v1.7.4.0917 (Bio-rad, Hercules CA) according to manufacturer’s directions. The fluorescence threshold was pre-set to 2000 RFUs. Reaction wells with <6,000 droplets were excluded. The HIV-1 LTR-gag concentration was estimated in copies per µL as the average of all evaluable wells, then multiplied by the well volume of 20 µL. The RPP30 concentration was corrected for dilution (1:10) and copies per cell (two per cell) to obtain number of cells per well, and total number of cells analyzed. HIV-1 LTR-gag copies per well was normalized to cells per well and expressed as HIV-1 LTR-gag copies per 10^6^ cells.

#### Determining assay performance

Analytic sensitivity and limit of detection (LoD), analytic specificity, precision and accuracy were determined (22) using diluted plasmids at the varying concentrations and with testing across two different blood donors in three independent runs. The limit of detection (LoD) was determined with a second set of experiments of 20 independent runs at the lower concentrations (5, 2.5 and 1.25 copies per 10^6^ cells). Validation panels were obtained from several sources. External validation for subtypes AE, AG, and C were provided by the Duke University External Quality Assurance Program Oversight Laboratory (EQAPOL); blinded samples (seven-three-fold dilutions of U1 cells (0 to 14,580 HIV-1 copies per 10^6^ cells spiked into PBMC pellets) from the NIAID Virology Quality Assurance (VQA) program for which four independent assay runs were conducted. The U1 cell line harbors two copies of HIV-1 genomic DNA per cell (23, 24), which was accounted for in the analysis. Serially diluted standards of HIV-1 subtypes A1, B and C diluted in one million uninfected cells) were provided by the National Institute for Biological Standard and Control (Potters Bar, United Kingdom). An additional dilution series was created with pellets of 5 million A3.01 cells spiked with increasing quantity of a mixture of two HIV-1 subtype A infectious molecular clones quantified by Nanodrop to examine lower sensitivity with subtype A standards spiked into one million cells.

#### Statistical Analyses

A least squares regression model was fit to the data using a simple linear model. The coefficient of determination, R^2^, was calculated to assess the linearity of this relationship overall and individually for each donor. Additionally, a second and third-order polynomial model was applied to the data. These models were then compared using an F-test to determine the model of best fit. A probit model, which uses maximum likelihood estimates to analyze a concentration-response relationship with a binomial response variable (detection or no detection by HIV-1 LTR-gag assay) was also performed to further assess the limit of detection (LoD) (25). Inter-assay variation was assessed by calculating the average measured HIV-LTR-gag copies per 10^6^ cells detected at each dilution level across the six independent assay runs, across two donors. Intra-assay variation was determined by assessing the variation across all six independent runs of the HIV-LTR-gag assay, across the two donors.

#### Study Approval

This study and use of Leukopaks from the NY Blood Center was approved by the Johns Hopkins Medicine Office of Human Subjects Research Institutional Review Boards as study number NA_00087629.

## Results

### Reportable Range and Linearity

A statistically significant linear relationship between expected plasmid concentration and measured HIV-1 LTR-gag copies (R^2^ =0.927 and p-value <0.001) was observed (Figure 1). With polynomial regression modeling and comparisons using F-tests, the data appeared nonlinear at the high end (1,000 copies per 10^6^ cells and above). Eliminating the highest analyte concentration (≥1000) showed that the data followed a linear model, with a slope significantly different from zero and a y-intercept not significantly different from zero (data not shown). The linearity of the assay at the highest concentrations across the range of 10,000, 1,000, 100 and 0 plasmid copies per 10^6^ PBMCs were retested across three independent runs. A statistically significant linear relationship between expected plasmid concentration per 10^6^ cells and measured HIV-1 LTR-gag copies (R^2^ =0.9968 and p-value <0.0001, Supplemental Figure 2) was found. With eight replicates of a 1000 ng of DNA, a median of 1,012,800 cells were analyzed (IQR 880,950 – 1,084,550).

**Figure 1.**
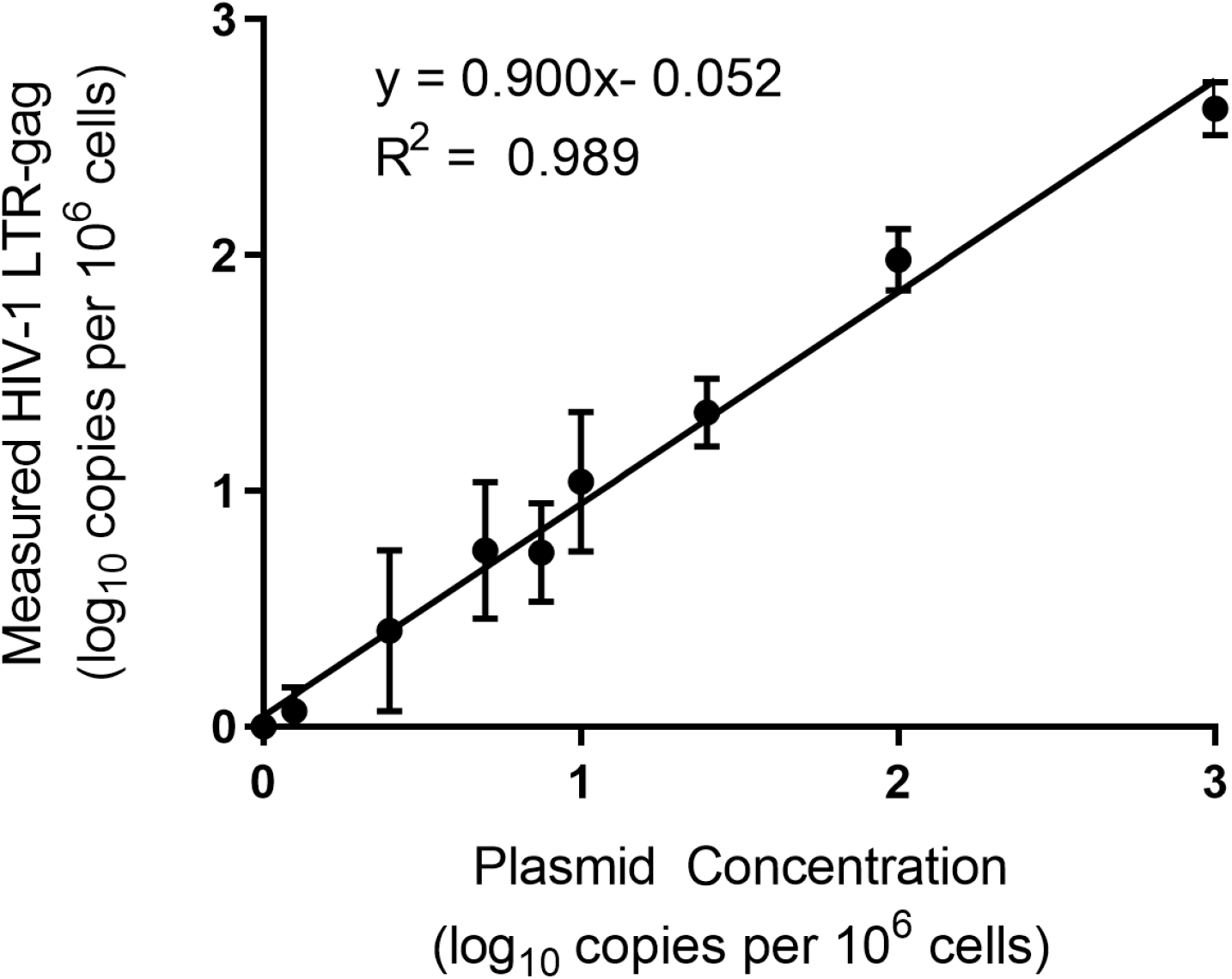
Measured HIV-1 LTR-gag copies (log_10_) from six replicate experiments with PBMCs from two different donors plotted against expected plasmid copies per 10^6^ cells (log_10_).

### Analytic Sensitivity

The assay was highly sensitive, detecting the presence of HIV-1 LTR-gag copies 100% of the time at concentrations of 1,000 to 5 copies per 10^6^ cells. The analytic sensitivity of the assay declined at 2.5 or lower HIV-1 DNA copies per 10^6^ cells, with approximately 83.33% and 33.33% detection at 2.5 and 1.25 copies per 10^6^ cells, respectively.

At the lower concentrations across 20 independent replicates, sensitivity of the assay was 100%, 60%, and 30% at 5, 2.5, and 1.25 copies per million cells, respectively. The LoD, the lowest concentration at which the assay can reliably detect 95% of the time, was estimated with the predicted probability line of 95% at 4.09 copies per 10^6^ cells assessed by the probit regression (Figure 2).

**Figure 2.**
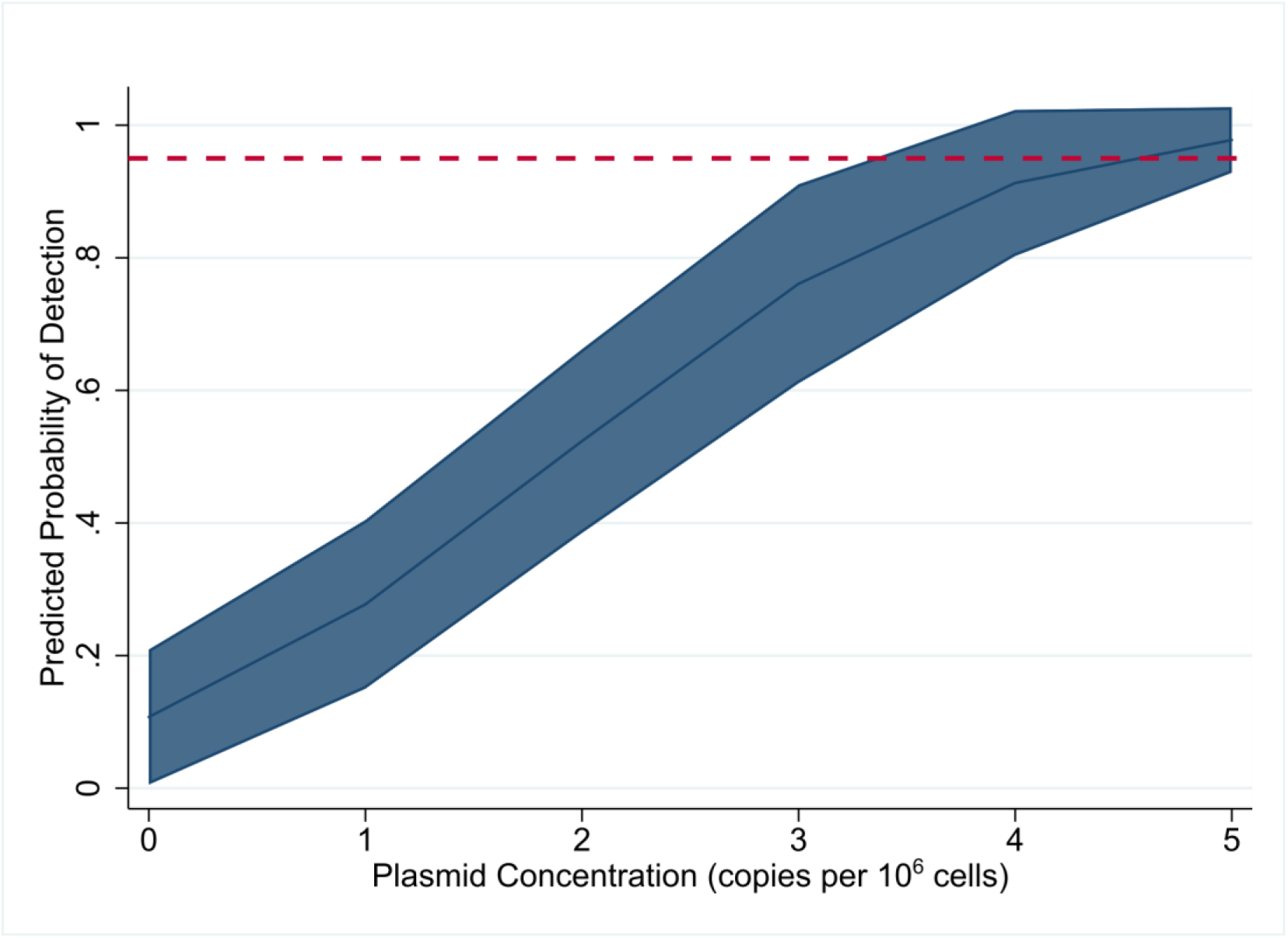
Predicted probability of detection by HIV-1 LTR-gag assay, as determined by probit analysis, at increasing plasmid copies per million cells with 95% confidence interval. Red dotted line depicts 95% probability of detection.

### Analytic Specificity

The diagnostic specificity of the HIV-1 LTR-gag assay was 100% with all six samples (three from each donor in three independent assay runs) without the plasmid DNA detected. Interference with hemoglobin was tested through analysis of blood samples saved as dried blood spots, no interference was detected (data not shown). Cross-reactivity to assess for the primer/probe to cross-react with genetically similar organisms was not performed.

### Inter-assay and intra-assay precision

The inter-assay standard deviation (SD) increased with the decreasing average concentration. At log_10_ 3 to 1.4 copies per 10^6^ cells, the SD range between 0.11 and 0.14; variation at the lower concentrations were higher at 0.30, 0.21 and 0.29 at log_10_ 1, 0.88 and 0.70 copies per 10^6^ cells, respectively (Table 1). The inter- and intra-assay %CV revealed a similar trend, with the lowest %CV across the two donors at 1,000 – 25 copies per 10^6^ cells (Supplemental Tables 2-3).

**Table 1.** Inter-assay precision

| **LOG<sub>10</sub> COPIES**<br>**PER MILLION**<br>**CELLS** | **LOG<sub>10</sub> AVERAGE**<br>**MEASURED HIV-1 LTR-**<br>**GAG PER 10<sup>6</sup> CELLS** | **STANDARD**<br>**DEVIATION** |
| --- | --- | --- |
| **3** | 2.62 (2.42 – 2.77) | 0.11 |
| **2** | 1.98 (1.83 – 2.13) | 0.13 |
| **1.40** | 1.33 (1.15 – 1.56) | 0.14 |
| **1** | 1.04 (0.64 – 1.31) | 0.30 |
| **0.88** | 0.74 (0.39 – 1.04) | 0.21 |
| **0.70** | 0.75 (0.21 – 0.98) | 0.29 |
| **0.40** | 0.41 (0 – 0.83) | 0.34 |
| **0.10** | 0.07 (0 – 0.20) | 0.10 |
| **0** | N/A | N/A |

### Accuracy

This analysis showed that the assay can accurately measure HIV-1 DNA at known input concentrations (Figure 3) with detection of approximately 92.82% of the expected copies per 10^6^ cells at each concentration level.

**Figure 3.**
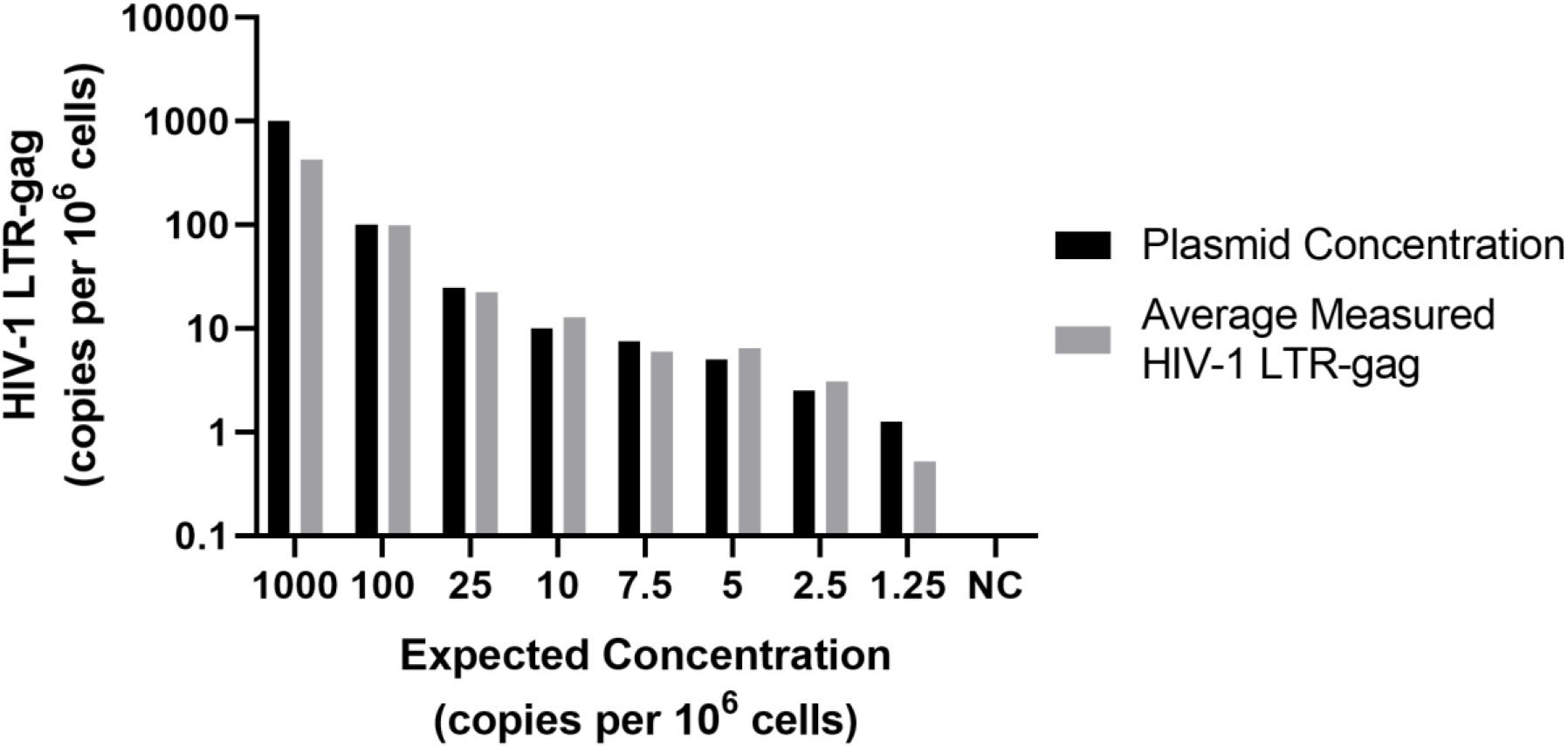
Comparison of expected plasmid (copies/million cells) and measured HIV-1 LTR-gag (copies per 10^6^ cells) to assess accuracy.

### External Validation

Testing of VQA program panel showed 100% detection of HIV-1 DNA at all concentrations (0, 60, 180, 540, 1,620, 4,860 and 14,580 copies per 10^6^ cells) in each of four independent runs and with no detection at zero copies HIV-1 DNA. The average measured HIV-LTR-gag log_10_ copies per 10^6^ cells detected and the degree of variation between all independent runs (%CV) with the imprecision around the four independent runs varying between 23.55% at the lowest concentration (60 copies per million cells) and 9.97% at the highest concentration (14,580 copies per 10^6^ cells). On a log scale, total imprecision (% CV) varied from 0.99% to 1.37%, with increased imprecision at lower concentrations with a 95% confidence interval of 1.21 to 1.58-fold change in measured HIV-LTR-gag (Table 2). The average percent detection at each concentration was higher with an overall quantification of the HIV-1-LTRGAG approximately 129.23% of the expected copies per 10^6^ cells, likely reflective of cell line standards harboring two proviral genomes per cell.

**Table 2.** Evaluation of precision in external validation samples by log %CV and 95% confidence interval of measured HIV-1-LTR-gag

| **DILUTION** | **LOG<sub>10</sub>**<br>**COPIES**<br>**PER 10<sup>6</sup>**<br>**CELLS** | **LOG<sub>10</sub>**<br>**AVERAGE**<br>**HIV-1 LTR-**<br>**GAG COPY**<br>**PER 10<sup>6</sup>**<br>**CELLS** | **STANDARD**<br>**DEVIATION** | **95% CI**<br>**(LOG<sub>10</sub> HIV-**<br>**1 LTR-GAG**<br>**COPY PER**<br>**10<sup>6</sup> CELLS)** | **LOG**<br>**CHANGE**<br>**95% CI** | **FOLD**<br>**CHANGE**<br>**95% CI** |
| --- | --- | --- | --- | --- | --- | --- |
| **VQA428 A** | 0 | 0 | 0 | N/A | N/A | N/A |
| **VQA428 B** | 1.78 | 1.99 | 1.36 | 1.88-2.08 | 0.20 | 1.58 |
| **VQA428 C** | 2.26 | 2.43 | 1.64 | 2.35-2.50 | 0.19 | 1.41 |
| **VQA428 D** | 2.73 | 2.84 | 2.14 | 2.74-2.92 | 0.18 | 1.51 |
| **VQA428 E** | 3.21 | 3.31 | 2.38 | 3.26-3.35 | 0.10 | 1.25 |
| **VQA428 F** | 3.69 | 3.74 | 2.80 | 3.69-3.78 | 0.10 | 1.25 |
| **VQA428 G** | 4.16 | 4.27 | 3.27 | 4.22-4.31 | 0.08 | 1.21 |

### HIV-1 Subtype analyses

HIV-1 subtypes AE and AG were readily detected using this assay, and displayed expected similar linear detection of at least 3-logs when tested on 10-fold serial dilutions of HIV-1 DNA from subtypes B and C spiked into 1 x10^6^ PBMCs, but not for subtype A (data not shown). However, when subtype A was reassessed with a preparation of 5 × 10^6^ million pellet that yielded sufficient genomic DNA to allow input of 8000 ng of genomic DNA, the performance and detection of subtype A was consistent with previous results for subtypes B and C (Figure 4A), and highly reproducible by two different operators (p <0.0001; Figure 4B).

**Figure 4.**
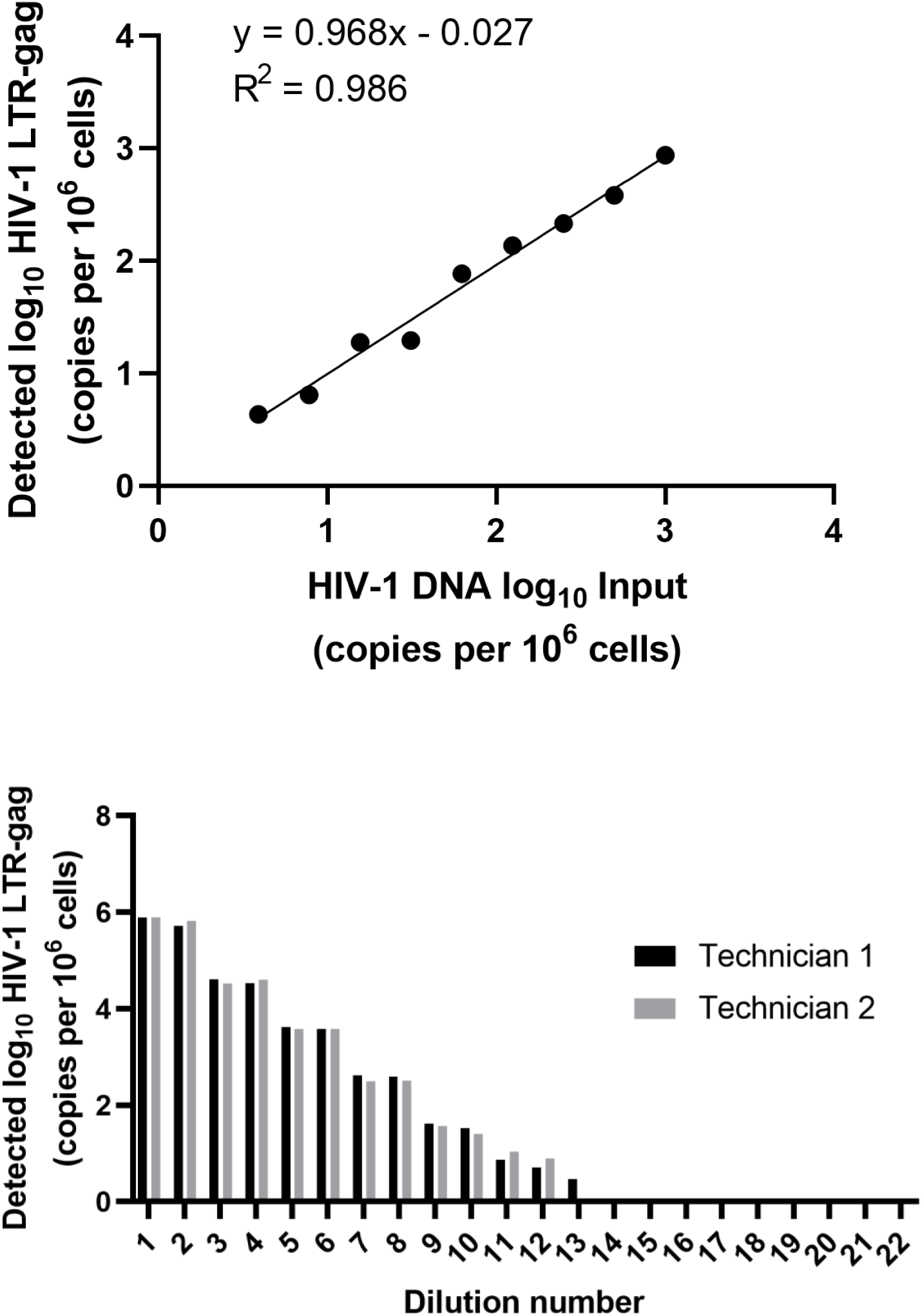
**A)** Measured copies of HIV-1 subtype A plotted against DNA spiked into uninfected cells. **B)** Variations in quantitation between two technicians processing samples in parallel.

## Discussion

ddPCR compared with qPCR employs the use of separation of the PCR mastermix into thousands of partitions, combined with Poisson statistics, which provides for precise and absolute quantification of a molecular target and minimizes interference with PCR inhibitors (19). By avoiding the need for a standard curve to infer target copy number as in qPCR, ddPCR leads to more reproducible and precise quantitation (22). However, discrepancies can occur with ddPCR when setting a threshold fluorescence at which a droplet is considered positive or negative. Variations in droplet fluorescence can stem from primer/ probe concentrations, baseline autofluorescence, and intermediate fluorescence referred to as rain that can complicate interpretation (26-28), Here, we found that setting a fixed threshold of fluorescence, we obtained highly reproducible, precise and accurate quantification of HIV-1 DNA present in samples with known input concentrations and across different HIV-1 subtypes.

The HIV-1 DNA assay reported here exhibits a statistically significant linear relationship between expected and measured HIV-1 LTR-gag copies with high diagnostic specificity, and reproducibility, and a lowest concentration of HIV-1 DNA detected as low as 1.25 copies per million cells with a predicted probability of LOD of 4.09 copies per million cells. This assay can effectively detect reproducibly across HIV-1 subtypes A, B, and C with equivalent performance characteristics, and with consistency in subtypes AE and AG. The eight replicates of 1,000 ng of genomic DNA input allowed an average of 1,034,258 cells to be analyzed, which likely contributed to the increased the sensitivity and accuracy.

Detecting and quantifying HIV-1infected cell concentrations is relevant to the fields of HIV-1 prevention and cure therapeutics (15, 17). Important insights into the immunopathogenesis of HIV-1 infection have been gained from studies of cell-associated infection during ART in infected individuals (29-33). In particular, early treatment of perinatal infection for which highly sensitive HIV-1 DNA assays become important for monitoring reservoir size. However, quantifying total HIV DNA with the ddPCR assay reported here is limited by its capacity to not distinguish intact from defective genomes as with the recently described intact proviral HIV-1 DNA assay (IPDA), which allows quantification of proviral genomes with potential to refuel infection (34, 35), although the sensitivity of the IPDA across a broad range of HIV subtypes has not been fully determined. Nevertheless, our HIV-1 LTR-gag assay has been shown to be sensitive and accurate across multiple HIV-1 subtypes allowing for assessment of low copy number samples without a priori knowledge of HIV subtype.

With enhanced HIV-1 prophylactic regimens that include combination antiretrovirals to prevent and empirically treat perinatal HIV-1 infection in high-risk exposure settings, access to more sensitive and precise HIV-1 DNA assays across various HIV-1 subtypes is becoming important. (16, 36).Similarly, the use of long-acting, potent antiretrovirals such as with long-acting injectables, including broadly neutralizing antibodies, to prevent adult infection, may alter the sensitivity of RNA and antibody testing, and for which ultrasensitive HIV-1 DNA testing may provide unique opportunities to detect occult, low level infection, enabling early transition to combination ART regimens to limit HIV-1 spread into reservoirs that preclude cure. In summary, this assay has potential applications for assessing therapeutic efficacy in the emerging fields of HIV-1 prevention, where occult HIV-1 infection may become more prevalent (37) and standard HIV-1 RNA and antibody testing methods insufficiently sensitive, and with relevance for HIV-1 cure, especially in perinatal infections, where HIV-1 infected cells concentrations can reach exceedingly low levels.

## Data Availability

No external datasets were used.

## Acknowledgements

We thank EQAPOL, the DAIDS VQA (HHSN272201200023C), the National Institute for Biological Standard and Control (Potters Bar, United Kingdom) for providing blinded samples for this study. U1 cells was obtained through the AIDS Research and Reference Reagent Program, Division of AIDS, NIAID, NIH: U1/HIV-1 from Dr. Thomas Folks. This research was supported by the National Institutes of Health (NIH) (R01 HD080474 (DP), PO1 AI131365 (DP)); the BELIEVE Collaboratory (1UM1AI26617); EPIICAL (16108367); the IMPAACT Center subspecialty laboratory (5UM1AI106716); UO1 (1U01AI135941) and the Johns Hopkins Center for AIDS Research (P30 AI094189).

